# Human genetic support improves the odds of drug success but poorly predicts it

**DOI:** 10.64898/2026.11.25.26368975

**Authors:** Leonhard Kohleick, Maik Pietzner

## Abstract

Nine in ten drug programmes that enter clinical trials fail, mostly for lack of efficacy^1,2^. Drug targets supported by human genetic evidence are about twice as likely to reach approval^3–5^, which now guides target selection. Yet most of it emerged only after the drugs had been developed, the methods linking genetic variants to genes were trained in part on known drug targets^6^, and recent trials of genetically supported targets have both lowered a disease marker^7^ and failed to reduce clinical events^8,9^. Here we tested how far genetic support can guide drug development across almost 17,000 target-disease pairs. Its association with clinical success held when genes were assigned without knowledge of drugs, within diseases and for evidence available years before a trial. It was strongest where the evidence pointed directly to the causal gene, larger for diseases judged on a measurable trait, and absent for drugs that use their target only to reach cells, such as antibody-drug conjugates. Genetic support, however, contributed little to distinguishing successful from failed programmes. Human genetic evidence is therefore a reliable but modest guide to where to invest, not a predictor of which programme will succeed.

## MAIN TEXT

Only about one in ten programmes entering clinical trials reaches approval^1,5^, and insufficient efficacy remains the most common, and costly, reason for failure in Phase II and III trials^2,10^. Naturally occurring genetic variation can provide evidence on the causal role of a target in human disease before any intervention is tested^11,12^. Genome-wide association studies (GWAS) provide the broadest and fastest-growing source of such evidence^13–17^. Whereas Mendelian evidence is largely restricted to rare diseases, GWAS span common diseases and the quantitative traits used as trial endpoints. Drug targets with human genetic support were about twice as likely to progress to approval^3^, independent of target and indication properties^4^, and a decade of further genetic discovery refined this to a 2.6-fold higher relative success that varied by therapy area, development phase and confidence in the causal gene^5^. Human genetic evidence has been proposed to support the majority of newly approved drugs^18,19^ and is integrated into public target-prioritisation resources^20^ and into models that forecast the outcome of drug programmes^21–23^.

Despite this widespread use, important gaps remain in how far genetic evidence can guide target selection (**Fig. 1a**). First, the reported association may be inflated by circularity and has not been re-examined as GWAS have grown. Most associated variants lie outside protein-coding regions, so the causal gene, and hence the putative drug target, has to be inferred^24,25^, commonly by variant-to-gene models trained in part on known drug targets^25^, and most genetic support for approved drugs emerged only after their development, with few targets genuinely discovered through human genetics^5,26,27^.

**Fig. 1.**
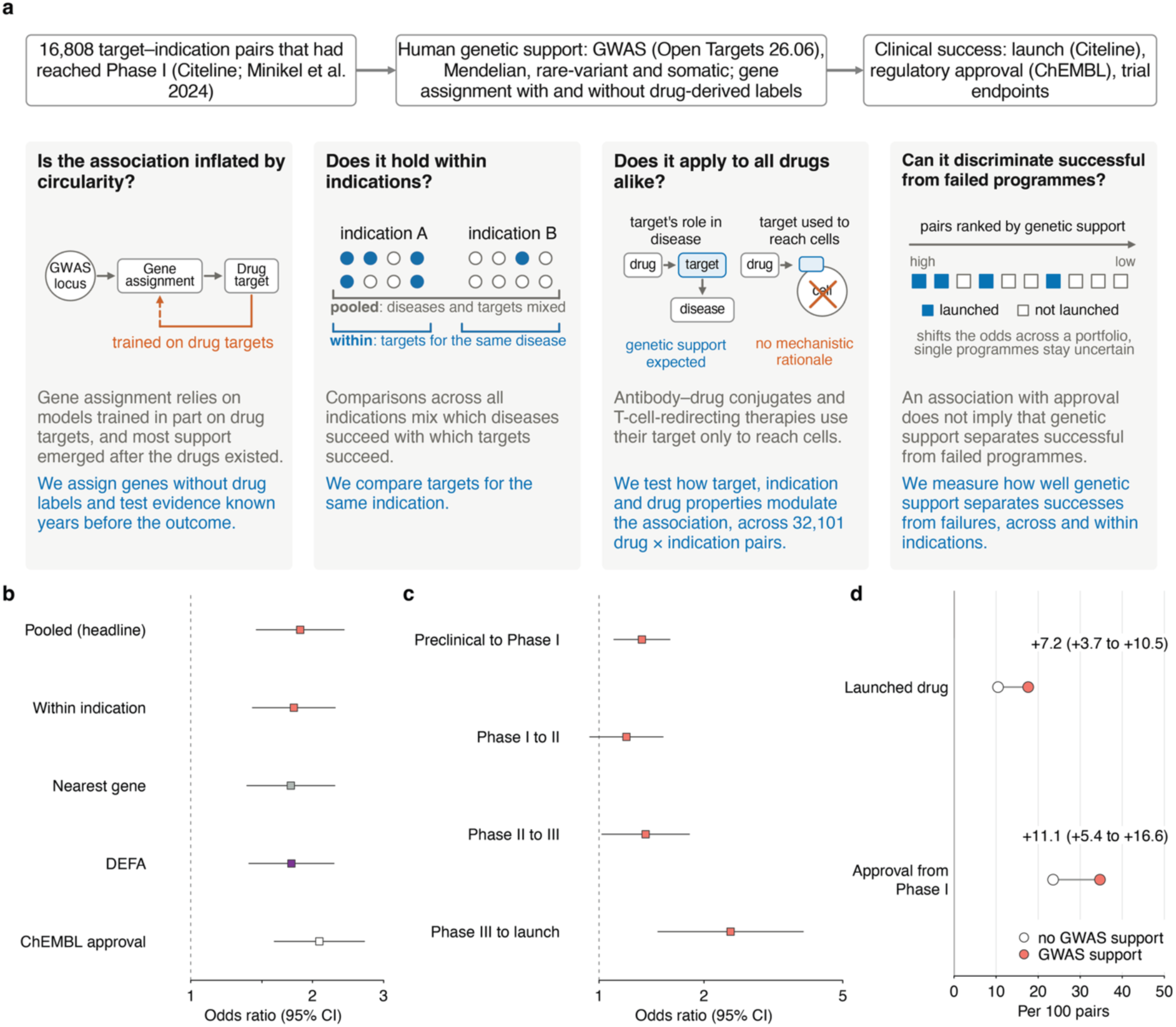
GWAS support and clinical success. **a**, Study design. We linked 16,808 target–indication pairs that had reached Phase I to human genetic support and to clinical success, and asked whether the association is inflated by circularity, holds within indications, applies to all drugs alike and discriminates successful from failed programmes. **b**, Odds ratios (ORs) for launch with GWAS support, overall, within indications, with two gene assignments that use no drug-derived labels (nearest protein-coding gene and DEFA), and for approval recorded in ChEMBL (open square). **c**, ORs for each phase transition. **d**, Launched drugs and approvals from Phase I entry per 100 pairs with and without GWAS support; numbers give the difference with its 95% confidence interval. Squares show ORs and lines 95% confidence intervals; the dashed line marks an OR of 1. Source data are provided as Source Data Fig. 1.

Second, the association has largely been treated as uniform across drugs. Its variation by therapy area and with properties of the genetic evidence has been characterised^4,5,28,29^, but not whether it depends on target biology, on whether success for an indication is judged on a quantitative trait that GWAS also measure, such as glycated haemoglobin (HbA1c) in diabetes, or on the mechanism of action. Antibody-drug conjugates and T-cell-redirecting therapies, for example, use their target only to reach the cells that express it, so there is no mechanistic rationale to expect genetic support for them. Likewise, evidence from rare Mendelian diseases and from GWAS loci linked to coding variants was more strongly associated with success than other GWAS evidence^4,5^, but the contributions of these sources have rarely been estimated jointly. Finally, an association with approval does not imply that genetic support can discriminate successful from failed programmes. Forecasting models of drug development rarely evaluated genetic evidence separately^21,22^, and recent models built on it were evaluated across indications^23,30^, which cannot separate the value of genetic support from differences in success rates between diseases.

Here, we linked the 16,808 target-indication pairs that had reached Phase I^5^ to current human genetic evidence spanning common, rare, and somatic variation and to gene assignments that use no drug-derived labels, including our own framework^31^, and evaluated success at the level of target-indication pairs, of individual trials and of 32,101 drug x indication pairs annotated by mechanism of action. We show that human genetic support is a stable but modest prior for drug development, whose value depends on the drug’s mechanism of action and on the endpoint, and that it does not reliably identify the programmes that will succeed.

### Updated GWAS support is robust to leakage and holds within diseases and in prospective tests

We first updated the association between GWAS support and clinical success for 16,808 target-indication pairs that had entered clinical development, of which 1,816 yielded a launched drug^5^. GWAS evidence from Open Targets (release 26.06) supported 773 pairs, which had almost twice the odds of yielding a launched drug (OR 1.86, 95% CI 1.45-2.40; **Fig. 1b**), after adjustment for therapy area, orphan status and gene pleiotropy, in line with earlier reports^3–5^. Only 147 of the 1,816 launched pairs had GWAS support and we explain **Supplementary Note 1** why most launched drugs lacked it. In absolute terms, 17.6% of supported compared with 10.4% of unsupported pairs yielded a launched drug, or 7.2 additional launched drugs per 100 pairs (**Fig. 1d**). We observed a similar estimate for regulatory approvals recorded in ChEMBL 36 (OR 2.08, 95% CI 1.61-2.69), and the association was strongest for the transition from Phase III to launch (OR 2.38, 95% CI 1.47-3.85) (**Fig. 1b,c**). Counting a pair as successful when at least two of three outcome sources agreed (see **Methods**) changed the outcome for 1,374 pairs but gave a similar estimate (OR 2.30, 95% CI 1.78-2.98 overall and 1.78, 95% CI 1.40-2.27 within indications). We found no evidence that well-studied targets or targets developed across many indications explained the association, neither when permuting support within genes (**Supplementary Tab. 1**) nor in a gene-level analysis (**Supplementary Tab. 1**).

Our updated support covered 225 more pairs than the most recent analysis (773 compared with 548)^5^, but only 337 pairs were supported in both. Expressed as relative success, GWAS support gave 1.83 (95% CI: 1.57-2.13) in our primary data set compared with 2.23 (95% CI: 1.90-2.62) when defined as by Minikel et al.^5^ for the same pairs (**Supplementary Tab. 1**). We deliberately did not count 52 pairs linked through measurement traits, such as triglyceride levels, as support (see **Methods**), and 31 pairs relied on studies no longer included in the current Open Targets release (**Supplementary Tab. 1**). The 436 pairs with support only in the current data launched less often (relative success: 1.45, 95% CI 1.16-1.83) (**Supplementary Tab. 1**), irrespective of when their support had been discovered (**Supplementary Tab. 1**), whereas counting pairs supported in either data set gave a similar overall estimate (OR 1.91, 95% CI 1.50-2.44) (**Supplementary Tab. 1**). Support that rested only on indirect trait mappings or on studies of several diseases at once was less informative, since counting such pairs as unsupported raised the estimate slightly (OR 2.02, 95% CI 1.52-2.67 and 1.96, 95% CI 1.50-2.56, respectively). More genetic support thus did not translate into more success per supported pair, emphasising the need to better understand for what types of drugs, their targets or indications the association holds.

The locus-to-gene (L2G) model, which assigns GWAS signals to genes, was trained in part on known disease genes, including targets of drugs that reached Phase III or approval^25^. About a quarter (24.2%) of supported pairs included an L2G training gene, 39.5% among launched compared with 20.6% among unlaunched pairs (**Supplementary Tab. 1**). Two gene assignments that use no drug or Mendelian disease labels, the nearest protein-coding gene and our DEFA framework^31^, corroborated the association in magnitude (OR 1.77 for both) (**Fig. 1b**; **Supplementary Tab. 1**), and launch rates increased with the confidence of the DEFA assignment (OR 1.13 per 0.1 increase in probability, 95% CI 1.05–1.22) (**Extended Data Fig. 1**). Half of the most confident assignments involved a coding variant, but the gradient remained among non-coding pairs (OR 1.11 per 0.1, 95% CI 1.02-1.19) (**Supplementary Tab. 1**).

At the level of individual trials, GWAS supported programmes tended to meet their primary endpoint more often once trial phase was taken into account (OR 1.24, 95% CI 0.94-1.64, and OR 1.29, 95% CI 1.01-1.64 after adjustment for tractability), whereas we observed no clear difference for stops due to lack of efficacy or safety (**Supplementary Tab. 1**).

Indications differed widely in how often their programmes succeeded, and supported pairs might simply have concentrated in diseases with high success rates. In a conditional logistic model stratified by indication, among 401 indications with both launched and failed programmes (n=14,894 pairs), GWAS support remained associated with success (OR 1.80, 95% CI 1.42-2.28), with similar estimates under both label-free gene assignments (**Fig. 1b; Supplementary Tab. 1**). GWAS support was therefore associated with the odds of success for a target chosen for a given disease and not just in general.

Most GWAS support for drugs was discovered after the drug programmes it was credited to. We therefore restricted analyses to support and phase entry documented by 2015, counting support discovered later as absent. GWAS support known in advance remained associated with progression from Phase I to II (OR 3.37, 95% CI 1.03-10.97), from Phase II to III (OR 1.97, 95% CI 1.06-3.67) and from Phase III to launch (OR 2.42, 95% CI 1.13-5.17) (**Fig. 2a; Supplementary Tab. 1**), although estimates were imprecise given the few pairs with support documented by 2015, and similar under label-free gene assignment (**Fig. 2a**).

**Fig. 2.**
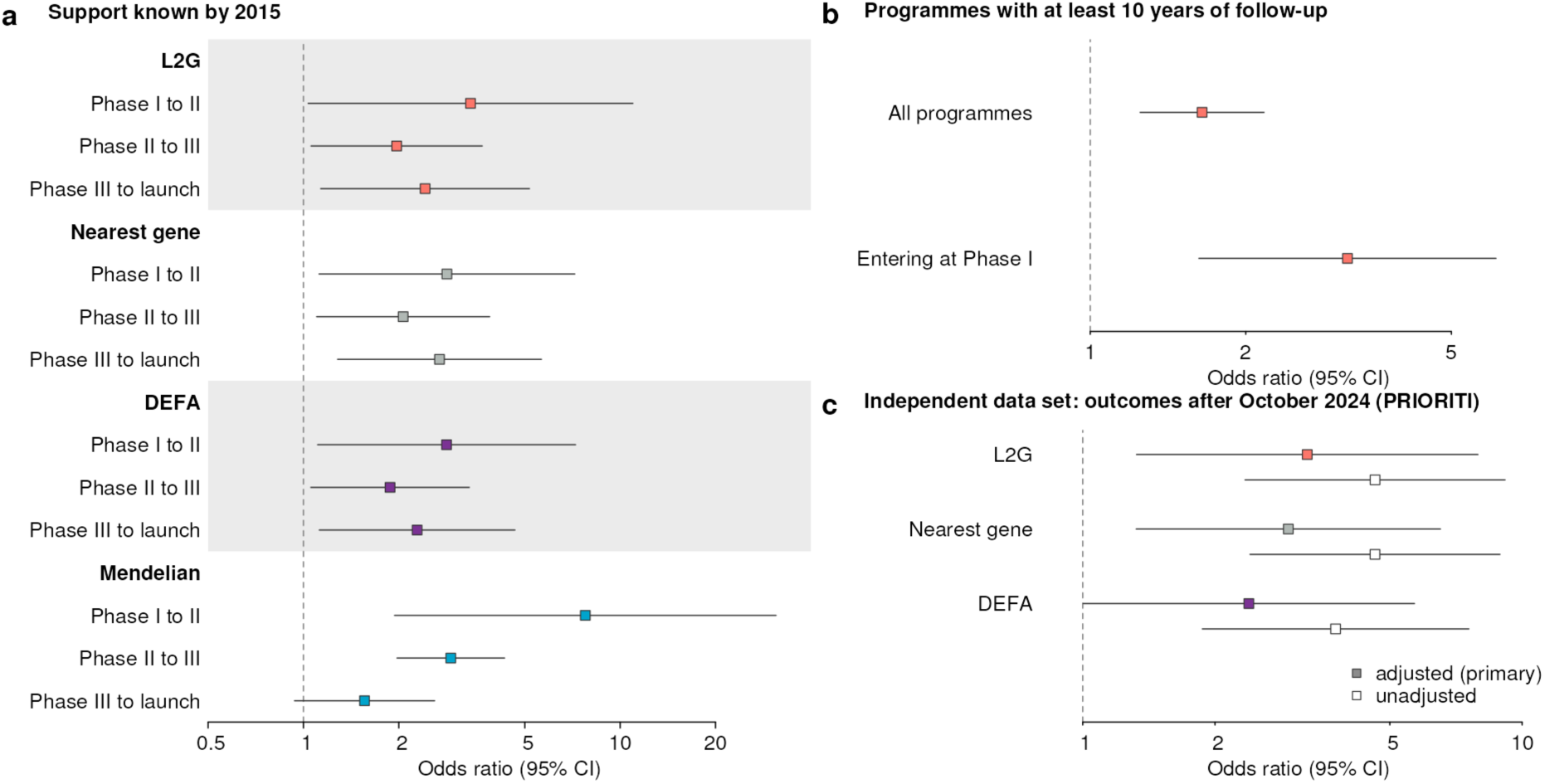
GWAS support known in advance. **a**, ORs for each phase transition when GWAS support and phase entry were documented by 2015, for three gene assignments (L2G, nearest protein-coding gene and DEFA) and for Mendelian support. **b**, ORs for programmes with at least ten years of follow-up (first recorded trial by 2012), for all such pairs and for those that entered at Phase I. **c**, ORs in an independent data set of 765 target–indication pairs whose outcome resolved after October 2024 (89 launched), using support documented by 2024; filled squares, adjusted for therapy area and pleiotropy; open squares, unadjusted. Colours indicate the gene assignment method (coral, L2G; grey-blue, nearest gene; violet, DEFA; cyan, Mendelian). Lines show 95% confidence intervals; the dashed line marks an OR of 1; log scale. Source data are provided as Source Data Fig. 2.

Programmes still ongoing when launch was recorded did not explain the association either. Among programmes that had entered trials by 2012, with at least ten years to resolve, the association was largely unchanged (OR 1.65, 95% CI 1.25-2.17) (**Fig. 2b**; **Supplementary Tab. 1**), and supported pairs were more likely to yield a launched drug whether ongoing programmes were counted as failures or assumed to resolve like completed ones (**Fig. 1d**).

The same held in an independent data set of 765 target-indication pairs whose development outcome resolved only after October 2024, compiled for a recent forecasting study^32^, of which 89 had launched. None of the 277 pairs also present in our primary data set had launched by its cutoff in December 2022. Using GWAS support documented by 2024 and adjusting for therapy area and pleiotropy, supported pairs were more likely to have launched than to have been discontinued (OR 2.94, 95% CI 1.32-6.52), with similar estimates under L2G and DEFA (**Fig. 2c**), with penalised estimation and when leaving out single genes (**Supplementary Tab. 1**). We note, that since the contrast excluded ongoing programmes, it is not directly comparable in size with the headline estimate reported above.

### The value of GWAS support varies with the drug and the endpoint more than with the target

We next asked whether the value of GWAS support depended on properties of the target, the indication, the drug or the genetic evidence itself. For each property, we estimated the odds ratio for GWAS support separately within each of its levels, for example among small molecules and among antibodies, and tested whether these odds ratios differed between levels by their ratio (an interaction). We judged each difference against robustness criteria set before the first such model was fitted (see **Methods**) (**Fig. 3**; **Supplementary Tab. 2**).

**Fig. 3.**
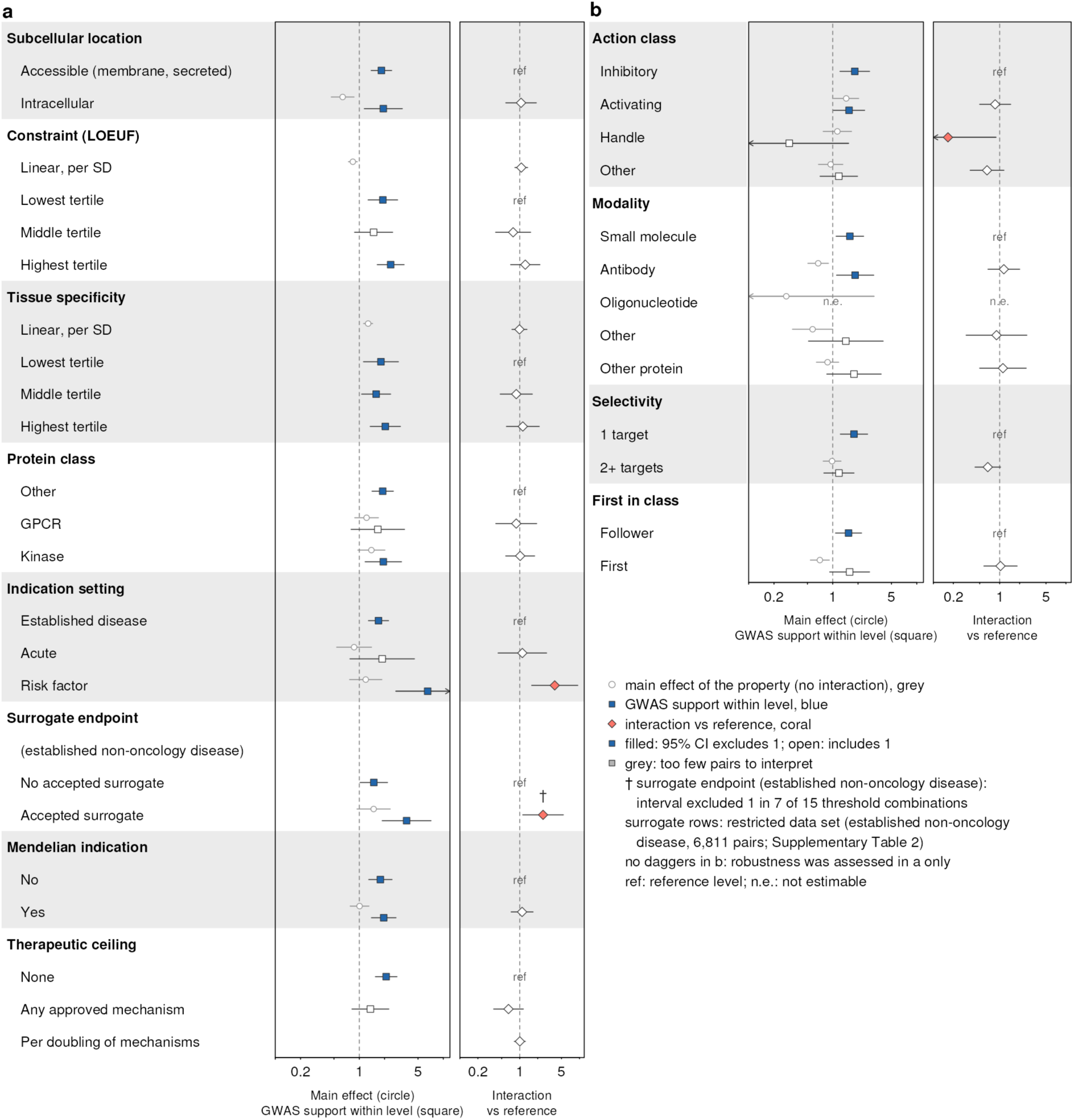
Target, indication and drug properties and the association of GWAS support with success. **a**, Target and indication properties among target–indication pairs, with launch as the outcome. **b**, Drug properties among 32,101 drug × indication pairs, with approval recorded in ChEMBL as the outcome. Left columns show the association of each property level with the outcome relative to its reference (grey circles; main effect) and the OR for GWAS support within each level (blue squares). Right columns show the ratio of the GWAS-support OR in each level to that in the reference level (interaction; coral diamonds). For constraint and tissue specificity, the linear rows give estimates per standard deviation; for the therapeutic ceiling, the rows give the step from no to any approved mechanism and the slope per doubling of approved mechanisms. The surrogate-endpoint rows are restricted to established non-oncology disease (6,811 pairs). Filled symbols, 95% confidence interval excludes 1; open symbols, interval includes 1; grey, levels with fewer than 200 pairs, 20 launches or 15 supported pairs. †, interaction that did not pass all robustness criteria (Methods). ref, reference level; n.e., not estimable. Log scale. Source data are provided as Source Data Fig. 3. Handle, drugs that use their target only to reach cells.

Several target properties were associated with the chance of launch individually, such as intracellular location (OR 0.63, 95% CI 0.46-0.87), genetic constraint (OR 0.84 per standard deviation, 95% CI 0.74-0.95) and tissue specificity (OR 1.27 per standard deviation, 95% CI 1.12-1.44), but none of them, nor protein class, robustly modified the association between GWAS support and success (interaction ORs between 0.88 and 1.06; non-linear codings p≥0.57) (**Fig. 3a**; **Supplementary Tab. 2**). Measures of the target’s *cis*-acting genetic architecture were neither associated with launch individually (none of five estimates excluded 1) nor modified the association (p=0.63) (**Extended Data Fig. 2**; **Supplementary Tab. 2**).

Observational molecular evidence added little. Differential expression of the target in the tissue affected by the disease was associated with success (OR 1.39, 95% CI 1.06-1.81), whereas targets whose plasma protein level predicted later onset of the indication in UK Biobank^33^ were not more likely to succeed (OR 0.93, 95% CI 0.66-1.31), and neither robustly modified the association with GWAS support (**Supplementary Tab. 2**).

We next asked whether GWAS support was more valuable for indications judged on a quantitative trait, which GWAS measure with high power, in two scenarios: the indication is itself a risk factor, such as hypercholesterolaemia or hypertension (see **Methods**), or an established disease with a quantitative surrogate endpoint accepted for approval, such as HbA1c in diabetes (**Fig. 3a**; **Supplementary Tab. 2**).

GWAS support was more strongly associated with success for risk-factor indications (OR 6.51, 95% CI 2.72-15.60) than for established disease (OR 1.69, 95% CI 1.28-2.23), and this difference kept its direction under ChEMBL approval, although its interval then included 1. In contrast, support among acute conditions did not differ from established disease (OR 1.87, 95% CI 0.77-4.54) (**Supplementary Tab. 2**). The risk-factor finding, however, largely applied to programmes without a recorded trial history and attenuated strongly among programmes with recorded trial dates (OR 2.13, 95% CI 0.91-4.95 within risk factors compared with 1.49, 95% CI 1.11-1.99 within established disease; **Supplementary Tab. 2**). It was driven, but not entirely explained, by lipid-lowering and antihypertensive targets (OR within risk factors without lipid indications: 4.94, 95% CI 2.35-10.36) (**Supplementary Tab. 2**), whose GWAS support was mostly discovered after approval, and therefore likely reflected genetics rediscovering the targets of drugs that had worked rather than predicting them.

To test the second scenario independent of risk factors (87% of risk-factor pairs also fell into indications with a surrogate endpoint), we restricted it to established, non-oncology disease. There, GWAS support was more strongly associated with success where a quantitative surrogate endpoint existed (OR 3.66, 95% CI 1.87-7.15 compared with 1.49, 95% CI 1.02-2.17 without), and this did not depend on programmes without recorded trial dates (**Supplementary Tab. 2**). This difference did not pass all robustness criteria, since its interval excluded 1 in 7 of 15 threshold combinations (**Supplementary Tab. 2**). The test was also not entirely independent of the outcome, because regulators accept surrogate endpoints partly because drugs acting on them succeeded, although restricting the analysis to surrogate endpoints in use before 2017 gave a similar result (OR 3.99, 95% CI 1.99-8.01 compared with 1.50, 95% CI 1.03-2.19). We therefore treat these results as hypothesis generating. Both scenarios nonetheless pointed to a greater value of GWAS support where success is judged on a quantitative trait (see **Discussion**).

Two further indication properties did not robustly modify the association (interaction ORs between 0.65 and 1.10) (**Fig. 3a**; **Supplementary Tab. 2**). We observed similar estimates in indications that were themselves monogenic diseases, and estimates did not clearly depend on how many mechanisms had already been approved for the indication when the programme began.

We tested drug properties in a separate data set of 32,101 drug × indication pairs with ChEMBL approval as the outcome, in which GWAS support was also associated with approval (OR 1.61, 95% CI 1.14-2.26). Drugs whose benefit does not depend on correcting the target’s role in the disease stood out, namely antibody–drug conjugates, T-cell-redirecting therapies, checkpoint inhibitors and lineage-depleting antibodies, for which the target serves to deliver a payload to, eliminate or engage particular cells. Such drugs almost never had GWAS support (1.4% of drug x indication pairs compared with 9.3% of other pairs), and among the 33 supported pairs only one, ipilimumab (targeting *CTLA4*) in melanoma, led to approval, so GWAS support showed no detectable association with approval for these drugs (interaction OR 0.24, 95% CI 0.03-0.96; **Fig. 3b**; **Supplementary Tab. 2**). Other drug properties, whether mechanisms were activating or inhibitory, antibodies or small molecules, directed at one or several targets, or first in class, showed no clear evidence for effect modification (interaction ORs between 0.66 and 1.16) (**Fig. 3b**; **Supplementary Tab. 2**).

We further extended GWAS support to proteins forming complexes or ligand-receptor pairs with the target^34–36^. This partner support was associated with success when derived from the genetic associations compiled by Minikel et al.^5^ (OR 1.30, 95% CI 1.04-1.62), but not when rebuilt from our 2026 GWAS support with the same interaction resource (OR 1.06, 95% CI 0.87-1.30), although the rebuild extended partner support from 1,970 to 3,048 pairs (**Supplementary Tab. 2**).

A recent analysis of Open Targets data reported that GWAS support through protein-altering variants, and support for genes of intermediate pleiotropy, were most predictive of approval^29^. We found no evidence that pleiotropy modified the association between GWAS support and success. Support through a coding variant, in contrast, was more strongly associated with success than non-coding support (OR 3.02, 95% CI 1.95-4.70 compared with 1.61, 95% CI 1.20-2.15), also when we rebuilt the earlier estimate step by step (ORs between 3.10 and 4.13) (**Supplementary Tab. 2**).

Taken together, the association between GWAS support and clinical success was stable across target biology, whereas we observed modifying effects by indication and drug class that require follow-up.

### Genetic evidence is most informative when it implicates the causal gene

Given that GWAS support through protein-altering variants was more strongly associated with success, we next added three further layers of human genetic support: Mendelian support from curated gene-disease resources, rare-variant support from gene-based burden tests, and somatic support from cancer driver catalogues^20^. With GWAS, Mendelian, rare-variant and somatic support mutually adjusted (the direct-evidence model), Mendelian support showed the strongest association with launch (OR 2.95, 95% CI 2.12-4.10), about twice that of GWAS support (OR 1.44, 95% CI 1.12-1.85). Somatic support was also associated with success (OR 1.84, 95% CI 1.19-2.85), whereas rare-variant support showed moderate enrichment but was estimated imprecisely (OR 1.56, 95% CI 0.90-2.71) (**Fig. 4a**). In absolute terms, 24.3% of pairs with Mendelian but no GWAS support yielded a launched drug, compared with 10.1% of pairs with neither.

**Fig. 4.**
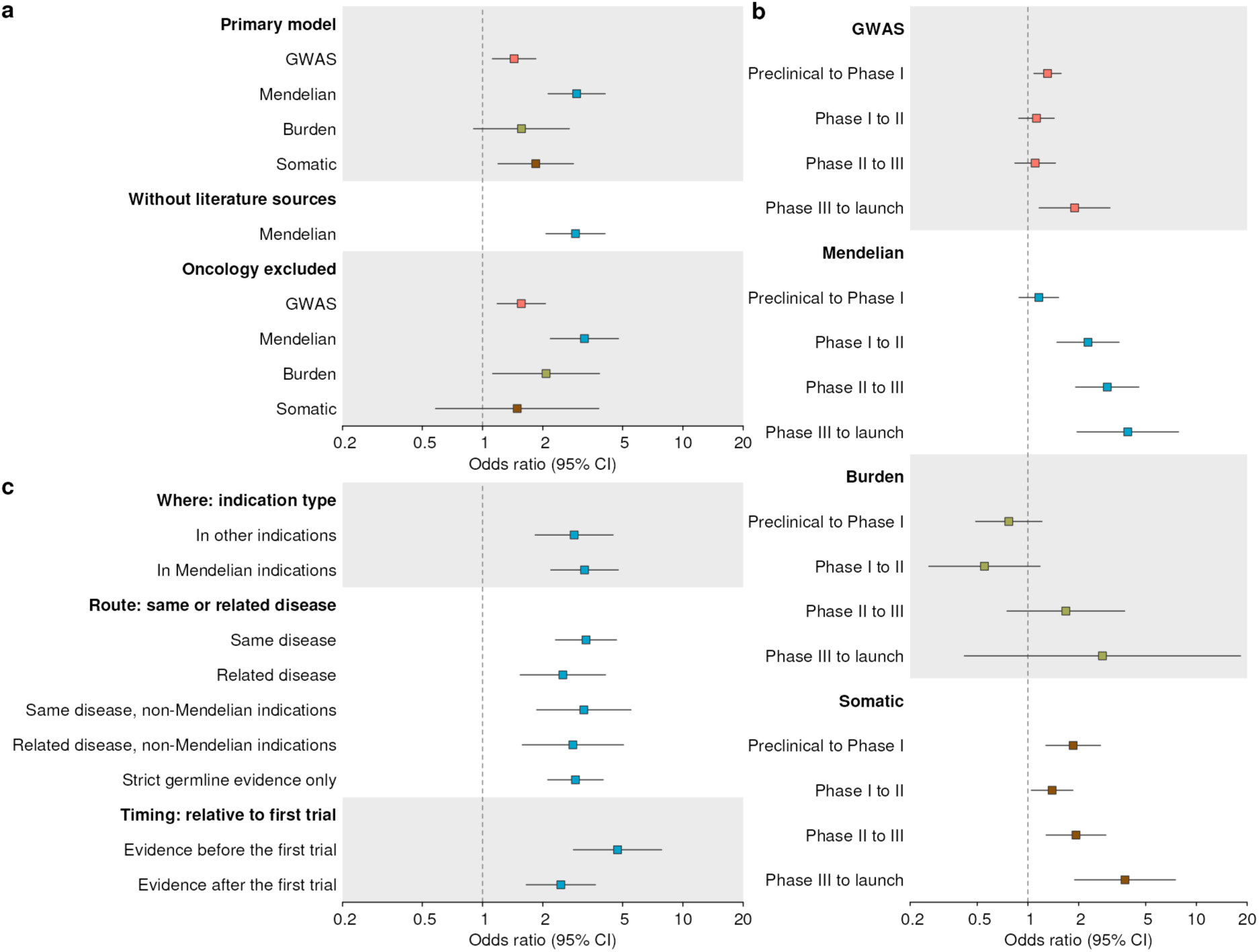
Sources of human genetic support. **a**, ORs for launch with GWAS, Mendelian, rare-variant (burden test) and somatic support in one model, and in two sensitivity analyses that excluded literature-curated Mendelian sources or oncology indications. **b**, The same four types of support for each phase transition; unlike Fig. 1c and Fig. 2a, these estimates use all support regardless of when it was discovered. **c**, Mendelian support split by indication type, by whether the evidence was recorded for the indication itself or for a related disease, and by whether it was available before the target’s first trial. Squares show ORs and lines 95% confidence intervals; the dashed line marks an OR of 1; log scale. Source data are provided as Source Data Fig. 4.

Mendelian support held within indications (OR 2.23, 95% CI 1.75-2.84) and under ChEMBL approval (OR 2.69, 95% CI 1.91-3.80), whereas the somatic estimate weakened under ChEMBL approval (OR 1.37, 95% CI 0.87-2.17) (**Tab. 1**). Like GWAS support, Mendelian support strengthened towards launch, from an OR of 1.16 before Phase I to 3.91 from Phase III to launch (**Fig. 4b**), and Mendelian support documented by 2015 was associated with later progression (**Fig. 2a**).

**Table 1.** Levels of evidence for the main observations of the present study. Each row is a central finding of this study and each column provides the evidence. Pairs (launched), the pairs that define the finding and how many yielded a launched drug; OR, launch, the primary estimate for launch recorded by Citeline; OR, approval, the same model for approval recorded in ChEMBL; label-free assignment, estimates with the nearest protein-coding gene and with DEFA. Four of the seven claims with estimates for both outcomes kept their conclusion under ChEMBL approval; partner support, estimated in two versions, is not counted. Values are ORs with 95% confidence intervals; interactions are ratios of ORs. The primary data set comprised 16,808 pairs, of which 1,816 yielded a launched drug and 1,446 an approval recorded in ChEMBL.

| ***Claim*** | ***Pairs (launched)*** | ***OR, launch (Citeline)*** | ***OR, approval (ChEMBL)*** | ***Label-free assignment, nearest gene / DEFA*** |
|---|---|---|---|---|
| *All Phase I+ target–indication pairs* | 16,808 (1,816) | – | 1,446 approved | – |
| *GWAS support* | 773 (147) | 1.86 (1.45-2.40) | 2.08 (1.61-2.69) | 1.77 (1.38-2.27) / 1.77 (1.39-2.26) |
| *GWAS support through coding variants* | 152 (45) | 3.02 (1.95-4.70) | 3.45 (2.25-5.29) | 3.25 (2.14-4.93) / 3.01 (1.94-4.67) |
| *Risk-factor vs established-disease indications (interaction)* | 205 (34) | 3.85 (1.58-9.38) | 2.23 (0.58-8.57) | 4.02 (1.75-9.25) / 3.68 (1.50-9.02) |
| *Quantitative surrogate endpoint (interaction)* | 845 (108) | 2.46 (1.12-5.38) | 1.69 (0.88-3.23) | 2.08 (0.94-4.60) / 1.89 (0.87-4.10) |
| *Handle drugs vs inhibitory drugs (Firth interaction, drug x indication data set)<sup>a</sup>* | 2,333 rows (193 approved) | 0.24 (0.03-0.96) | – | – |
| *Partner support (2024 associations; 2026 rebuild)<sup>b</sup>* | 1,970 (279) | 2024: 1.30 (1.04-1.62); 2026 rebuild: 1.06 (0.87-1.30) | 2024: 1.77 (1.45-2.16) | 2026: 1.09 (0.90-1.32) / 0.98 (0.81-1.18) |
| *Mendelian support (direct-evidence model)* | 447 (149) | 2.95 (2.12-4.10) | 2.69 (1.91-3.80) | – |
| *Burden-test support (direct-evidence model)* | 83 (31) | 1.56 (0.90-2.71) | 1.50 (0.84-2.68) | – |
| *Somatic support (direct-evidence model)* | 1,543 (220) | 1.84 (1.19-2.85) | 1.37 (0.87-2.17) | – |

<sup>a</sup> Rests on one approval among 33 supported drug x indication pairs. <sup>b</sup> The association in the 2026 rebuild did not replicate the 2024 estimate.

Three ablation analyses, each compared with the direct-evidence estimate (OR 2.95), showed that the Mendelian association did not depend on rare-disease indications. Mendelian support was associated with success both in indications that were themselves monogenic diseases, such as cystic fibrosis or haemophilia A (OR 3.23, 95% CI 2.19-4.76), and in all other indications (OR 2.87, 95% CI 1.83-4.49). The evidence also did not need to be recorded for the indication itself. Records for the indication itself and records for a rarer disease that our crosswalk linked to it gave similar estimates in non-monogenic indications (OR 3.20, 95% CI 1.86-5.51 and OR 2.82, 95% CI 1.58-5.06). Finally, defining Mendelian support by germline evidence alone gave a similar estimate to the definition including literature-derived sources (OR 2.91, 95% CI 2.11-4.00) (**Supplementary Tab. 3**).

Mendelian evidence that predated the target’s first clinical trial was more strongly associated with success (OR 4.72, 95% CI 2.84-7.84) than evidence discovered later or undated (OR 2.46, 95% CI 1.65-3.66). This argued against the possibility that drug development itself had generated the Mendelian evidence (**Supplementary Tab. 3**).

The ambiguous association with rare-variant support from burden testing warrants a note, given recent large-scale investments in exome sequencing of population cohorts. Such support was available for only 83 pairs, which were associated with success on their own (OR 2.55, 95% CI 1.40-4.65) but largely overlapped with Mendelian support (OR 1.56, 95% CI 0.90-2.71), since only 36 burden-supported pairs lacked Mendelian support, with 4 launches among them (**Supplementary Tab. 3**). An observation in line with population-scale exome studies in which most gene-based associations with disease involved genes already known from Mendelian disease^37^.

### Genetic support weakly separates successful from failed programmes

We then asked how well human genetic support (GWAS, Mendelian, rare-variant, somatic and risk-factor-trait support), alone or with target biology, could discriminate successful drug programmes, strictly splitting out-of-fold evaluations by target, so that no target appeared in both training and testing. We measured discrimination as the area under the receiver operating characteristic curve (AUC) with values of 0.5 being chance only. A base model containing only the therapy area of each indication reached an AUC of 0.573. Target biology added more discrimination than genetic support on its own (AUC 0.643 compared with 0.613), and both together reached 0.662 (**Fig. 5a**; **Supplementary Tab. 4**). Much of the biological signal, however, was shared with other target annotations, such as tractability and annotation completeness. Selectively removing target biology from the full model led to a similar decrease in AUC (0.022) as omitting genetic support (0.017), whose contribution was barely shared with any other annotation. The biological increment (the gain in AUC from adding target biology to the therapy-area model) was unchanged when we removed annotations that partly record whether a target had already been drugged (0.072 compared with 0.070) (**Supplementary Tab. 4**). Other machine learning models did not do materially better (AUCs between 0.622 and 0.669), with Mendelian support the only genetic term among the largest LASSO coefficients (**Extended Data Fig. 3**). The genetic association score of Open Targets, built from the same sources, added nothing beyond our genetic layers (within-indication increment: 0.001), and our layers little beyond it (0.008) (**Supplementary Tab. 4**).

**Fig. 5.**
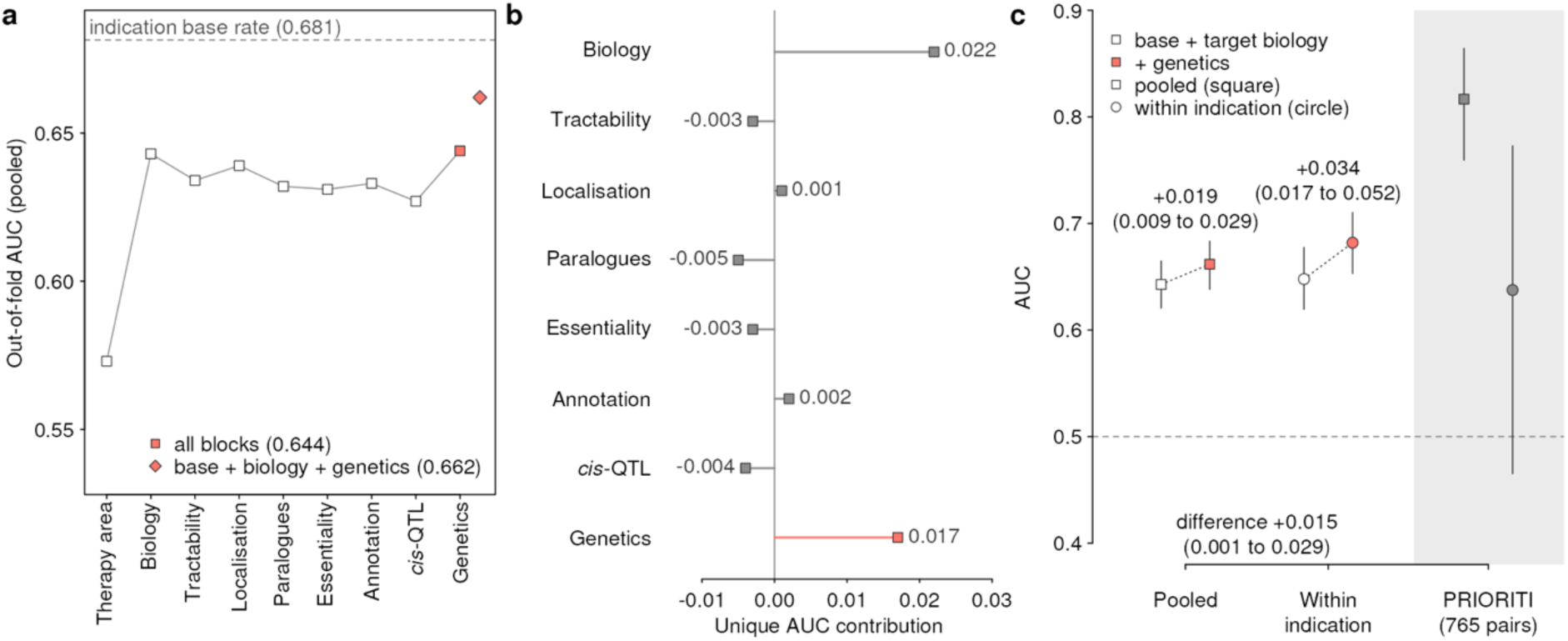
Discrimination between successful and failed programmes. **a**, Area under the receiver operating characteristic curve (AUC) as blocks of predictors were added in a fixed order, from out-of-fold predictions with folds grouped by gene; square, all blocks; diamond, therapy area, target biology and genetic support; dashed line, a model using only each indication’s historical success rate. **b**, Loss in AUC when each block was left out of the full model. **c**, Pooled and within-indication AUC for therapy area and target biology (open symbols) and after adding genetic support (filled symbols), with squares for pooled and circles for within-indication estimates, with the gain and its 95% confidence interval from 2,000 bootstrap samples of indications; for comparison, the published predictions of PRIORITI^23^ on its own data set of 765 pairs (shaded). Source data are provided as Source Data Fig. 5.

Machine-learning models built to forecast drug development have reported AUCs of 0.78 to 0.88^21,22^, mostly using features, such as trial results or sponsor track record, that become available only after a target has been chosen. Much of such a pooled AUC can come from knowing which indications tend to succeed. A model using only each indication’s historical success rate reached a pooled AUC of 0.681, higher than our full model, although by construction it cannot rank targets for the same disease. Within indications, our model reached an AUC of 0.682 (95% CI: 0.653-0.710), as high as its pooled AUC, and adding genetic support yielded a greater increase than in the pooled scenario (0.034 compared with 0.019; difference: 0.015, 95% CI 0.001-0.029) (**Supplementary Tab. 4**). In contrast, the published predictions of a recent forecasting model built on large language models fell from an AUC of 0.817 pooled to 0.638 (95% CI: 0.465-0.773) within indications on its own post-2024 data set^23^ (**Fig. 5c**).

We note that persistent associations and weak discrimination are not mutually exclusive. In the post-2024 data set, GWAS support was strongly associated with launch (OR 2.94, 95% CI 1.32-6.52), yet ranked pairs with an AUC of only 0.547 (95% CI: 0.513-0.590), because only 31 of the 765 pairs carried support, and about 14 target-indication pairs needed GWAS support for one additional launched drug (**Fig. 1d**; **Supplementary Tab. 1** and **4**). Human genetic support therefore shifted the odds of success across a portfolio, but neither it nor target biology could reliably identify the programmes that would succeed. This was partly expected, since 67% of stopped trials with a recorded reason in our trial-level data set stopped for reasons unrelated to the therapeutic hypothesis, such as recruitment or business decisions, in line with earlier work^38^, and GWAS support was not associated with these stops (OR 0.96, 95% CI 0.81-1.13) (**Supplementary Tab. 4**).

## Discussion

Human genetic support has been widely adopted for selecting drug targets, largely because of retrospective comparisons of drug programmes across all diseases. Here, we tested whether this association holds when genes are assigned without drug-derived labels, within diseases and for all drugs alike, and whether it translates into prediction of success. GWAS support remained associated with clinical success within diseases, prospectively, in programmes that resolved after 2024 and with gene assignments free of drug-derived labels. The association was more modest than previously estimated, did not depend on properties of the target, such as subcellular location, genetic constraint or protein class, differed between types of drugs and was stronger for indications judged on a quantitative trait. Genetic support, alone or with target biology, could not reliably discriminate between successful and failed drug programmes.

Minikel et al. found that the value of GWAS evidence did not decline with its year of discovery and concluded that genetic insights for drug discovery are far from saturated^5^. Recently discovered support was not less informative. However, support identified only in the current data was less strongly associated with success than support present in both the earlier and the current data. Partner support, which extends GWAS evidence to interacting proteins, reached many more pairs when rebuilt from current data but was no longer associated with success. As GWAS grow, more genes likely gain support for some disease, and the information carried by any single link may fall unless the evidence points specifically to the causal gene. Coding variants and Mendelian disease, which provide the clearest causal gene evidence, were more strongly associated with success throughout, as were rare, large-effect associations and confident gene mapping in an independent cohort^28^, which suggests weighting genetic support by its specificity rather than counting it. Evidence at non-coding loci was less informative, and whether this reflects uncertain gene assignment or regulatory effects that drugs and model organisms capture poorly remains open.

Genetic support is commonly read as a property of the target that benefits any drug acting on it, and Minikel et al. hypothesised that its value depends on how a drug acts but lacked the data to test this^5^. We identified a group of drugs for which genetic support carried no detectable information, namely antibody-drug conjugates, T-cell-redirecting therapies, checkpoint inhibitors and lineage-depleting antibodies. These drugs use their target only to reach particular cells rather than to correct its role in disease, natural variation in such targets has no reason to influence disease risk, and they rarely had genetic support. Because these classes account for a growing share of approvals, particularly in oncology^39^, ranking such targets by genetic support would likely falsely deprioritise them, and estimates pooled over all drugs understate the value of genetic support for drugs that act through the target’s role in disease.

GWAS support was more strongly associated with success where success is judged on quantitative traits that are also studied by GWAS, for indications that are themselves risk factors and for diseases with a surrogate endpoint accepted by regulators. The same held for the pairs linked through measurement traits that distinguished the earlier work from ours, which often involved drugs approved on the very trait that GWAS measured, such as thrombopoietin-receptor agonists (*MPL*) for thrombocytopenia, for which the genetic evidence emerged only after launch. None of the 25 pairs linked through such a defining measure had genetic evidence before its first trial (**Supplementary Tab. 1**). GWAS of quantitative traits are better powered and measure biology closer to the endpoint on which approval rests, and genetics may have rediscovered the targets of drugs long in use, as the attenuation of the risk-factor finding among programmes with recorded trial dates suggests. Where approval rests on the trait itself, genetic support may predict regulatory success almost by construction but says less about benefit to patients. For example, the CETP inhibitor obicetrapib was approved in Europe in 2026 on its lowering of low-density lipoprotein cholesterol, with its cardiovascular outcome trial yet to report^40–42^, and silencing *CIDEB*, a target identified through rare coding variants^43^, lowered liver fat in a first trial^7^ but effects on liver histology and outcomes remain to be shown.

Our results also help to understand how the two prominent failures of 2026 could be interpreted. Ziltivekimab, an antibody against interleukin-6, and pelacarsen, an antisense oligonucleotide against apolipoprotein(a), both lowered their targets markedly but did not reduce cardiovascular events in large outcome trials^8,9^. Pelacarsen rested on coding-variant evidence in the target gene^44^, which our analysis ranked among the most informative, whereas for ziltivekimab the coding evidence lies in the receptor gene *IL6R*^45^ and reaches the drug’s target only as partner support, which did not replicate in our data. Both were tested on hard cardiovascular outcomes in established disease, the setting in which genetic support was least informative (**Supplementary Tab. 2**), and in patients already on guideline-directed therapy, where lowering the target late in life may achieve less than the lifelong difference in exposure that genetic variants confer as noted by authors of the HORIZON trial^9^. Neither trial, however, can be explained by an aggregate analysis, and each may have failed for its own reasons, but neither is surprising given our results. From Phase III entry, programmes with coding-variant support reached launch in about three of four cases, whereas in cardiovascular indications supported programmes launched in only about one of two, hardly more often than unsupported ones.

Association with success did not translate into the ability to identify successful programmes. Genetic support improved the odds of success across a portfolio, but neither it nor target biology separated successful from failed programmes well, and evaluations across diseases largely measured which diseases rather than targets tend to succeed. Observational molecular evidence is unlikely to close this gap, since differential expression in diseased tissue was less strongly associated with success than genetic support (OR 1.39 compared with 1.86) and plasma proteins that prospectively associated with disease onset were not associated with success at all (OR 0.93). A model using only each indication’s historical success rate performed as well as our full model, and a recent LLM-based forecasting model lost most of its performance within indications, where the contribution of genetic support was in fact larger. Forecasting models should therefore report their performance within indications, the comparison that matches the choice between targets for a given disease. Genetic support was also not associated with trial stops for lack of efficacy, although insufficient efficacy is the most common reason for failure in Phase II and III^2^. In the terms of decision theory, genetic support is a decision tool of modest predictive validity^46^, and because successful programmes are rare, even modest predictive validity improves the yield of a portfolio. Genetic support is thus better suited to guide decisions across a portfolio than to predict the outcome of single programmes.

Our study is distinguished by evaluating genetic support within diseases, prospectively and without leakage from gene assignment, but several limitations need to be considered. Firstly, the outcome data had been analysed in earlier versions of this work, so we present all analyses as descriptive rather than confirmatory and we documented each specification and when it was set in the code repository. Secondly, launch was recorded up to December 2022, so programmes still ongoing counted as not launched, although results were similar for mature programmes and for approvals recorded in ChEMBL. Thirdly, genetic support depended on how GWAS traits were mapped to indications, our mapping did not treat measurement traits as disease equivalent, and two-thirds of the credible sets behind GWAS support came from studies of European ancestry only, although 58% of supported pairs also had support from multi-ancestry or non-European studies (**Supplementary Tab. 1**). Fourthly, our primary outcome derives from a single proprietary database with Western regulatory histories, whose launch records agreed with regulatory approvals for fewer than half of all launches (844 of 1,816), and absolute estimates are lower in broader open registries^47^. Finally, findings on drug classes and indication settings rested on few events and should be read as hypotheses, we had limited power to detect small modifications, and our models used public annotations only. Two in five launched pairs with GWAS support included a gene in the L2G training set, yet gene assignments free of drug-derived labels gave the same estimate, but we recommend comparing rankings of new targets against such label-free assignments.

In conclusion, human genetic support is a stable but modest prior for drug development, with about 14 target-indication pairs needing GWAS support for one additional launched drug. Its value depends on the drug’s mechanism of action and on the endpoint on which approval rests, and lies in guiding where to invest rather than in predicting which programme will succeed. In practice, this means weighting genetic support by its specificity and judging drugs that use their target only to reach cells via other means. Our findings also imply that forecasting models must be evaluated within and not across indications.

## Methods

### Study design and outcomes

We used the target-indication pairs assembled by Minikel et al.^5^ from drug programmes in Citeline Pharmaprojects (snapshot of 22 December 2022), restricted to the 16,808 pairs that had reached Phase I, spanning 1,603 targets and 876 MeSH indications. The primary outcome was the launch of a drug for the pair, as recorded by Citeline (1,816 pairs). As a second outcome, a pair counted as approved if a drug acting on the target had reached phase 4 for the same indication in ChEMBL 36 (1,446 pairs). Phase transitions followed Minikel et al.^5^. The phase of a pair was the highest phase reached by its programmes if none was active, and otherwise the phase of its active programmes, and a pair counted as launched if any drug for the pair had launched. In a sensitivity analysis, we counted a pair as successful if at least two of three sources agreed (launch in Citeline, approval in ChEMBL and approval in Open Targets, whose clinical records draw in part on ChEMBL).

### Human genetic support

We counted a pair as supported by GWAS if a credible set in Open Targets 26.06 assigned the target with an L2G score of at least 0.5 for a study trait that was either the indication itself or had a similarity of at least 0.8 to it, using the similarity measure of Minikel et al. ^5^. We used no other relations between traits and indications. We mapped study traits recorded in disease ontologies (EFO, MONDO, HP, Orphanet or DOID) to MeSH indications through Open Targets cross-references. A trait without a cross-reference of its own took that of its parent or child term, one step away in the ontology, with at most 20 inherited terms per MeSH term. We did not map traits from other ontologies, such as Gene Ontology terms. Because L2G was trained in part on drug targets^25^, we repeated key analyses with two gene assignments that use no drug-derived labels (label-free), namely the gene carrying a protein-altering variant in the credible set (posterior inclusion probability ≥ 0.1), or else the nearest protein-coding gene, and the highest-scoring gene of DEFA^31^ in its label-free model set, which excludes the classifier trained on Open Targets gold-standard genes. We matched genes to Open Targets targets by approved symbol and did not remap ten target genes recorded under a superseded symbol. Matching them would have added GWAS support to three Phase I+ pairs and Mendelian, burden or partner support to six, one of them launched, without changing any conclusion. To quantify leakage, we matched supporting credible sets to the L2G training set by study locus, lead variant or trait. We took the ancestry of each supporting study from the discovery sample recorded in the Open Targets study table.

We used curated germline evidence to capture Mendelian support when linking the target to the indication (Orphanet, Gene2Phenotype, ClinGen, Genomics England PanelApp, ClinVar and UniProt, with inclusion levels in **Supplementary Tab. 5**). For rare-variant support, we used gene-burden associations, and for somatic support cancer-driver evidence, all from Open Targets and mapped to indications at the same similarity threshold. We counted Mendelian support as predating the first trial of the target if its earliest dated evidence preceded the target’s first recorded trial in any indication. Because L2G was trained in part on genes from the same Mendelian resources^25^, we mutually adjusted for GWAS and Mendelian support to account for their overlap.

We manually linked 13 clinical traits (cholesterol, glucose, urate, calcium, iron and red cells, white cells and inflammation, eosinophils, platelets, C-reactive protein, Immunoglobulin E, van Willebrand Factor, body mass index and blood pressure) to the indications they predispose to, augmenting and refining earlier work^29^, and counted a target as having risk-factor-trait support when it had GWAS support (credible set, L2G ≥ 0.5) for a linked trait. Where the trait credible set colocalised (H4 ≥ 0.70) with a credible set from a GWAS of the pair’s own indication, we checked whether the allele’s effect on the trait, in the direction of the drug’s action, matched its effect on the disease, and classed each supported pair as for, against, untested or not testable (**Supplementary Tab. 5**). We note, that this layer covers traits upstream of a separate disease and not disorders defined by the trait itself, such as hypertriglyceridaemia for triglyceride levels.

To define partner support, we extended genetic support to interaction partners of the target, from protein complexes of up to 25 proteins, ligand-receptor pairs and curated interactions. The 2024 term used the genetic associations compiled by Minikel et al.^5^. Our rebuild used the 2026 GWAS support of the partner genes (L2G), or any of the four direct layers, and we repeated it under both label-free assignments. We added each version to the layers model (see below), overall and separately within and outside oncology.

### Statistical analysis

For the headline estimate and its sensitivity analyses, the analyses of modifiers and the absolute estimates, we fitted the GWAS-only model, a logistic regression of the outcome on GWAS support adjusted for pleiotropy, therapy area and orphan status, with standard errors clustered two ways, by gene and by indication (CR1 sandwich estimator). We defined pleiotropy as the number of MeSH indications with GWAS support for the gene, in five bins (0, 1, 2-3, 4-7, 8+), computed for each gene assignment strategy separately. We fitted two further models. The direct-evidence model included GWAS, Mendelian, rare-variant and somatic support as separate, mutually adjusted terms. The layers model added the four classes of risk-factor-trait support to the direct-evidence model and served the partner-support sensitivity analysis. We used the genetic associations compiled by Minikel et al.^5^ only for the 2024 partner term, for the comparison with their definition of GWAS support and for dating support in the prospective analyses. For the drug x indication data set we adjusted for pleiotropy and therapy area (see below), and for trial-level models we added trial phase to the covariates of the GWAS-only model, with tractability covariates as a sensitivity analysis.

For sensitivity analyses of the headline GWAS support estimate, we reassigned support 1,000 times within each gene, keeping each gene’s number of supported indications and sampling indications in proportion to their frequency in the primary data set. For a gene-level estimate, we sampled one pair per gene 500 times and averaged the odds ratios of the GWAS-only model. Within indications, we fitted conditional logistic regression models stratified by indication (Efron approximation), with GWAS support and pleiotropy, and standard errors clustered by gene. To allow programmes time to resolve, we repeated the analysis in pairs whose first recorded trial started in 2012 or earlier, giving at least ten years of follow-up. In a further sensitivity analysis, we counted pairs supported in either the earlier or the current data as supported.

In the prospective analyses, we counted support as known in a given year if its first-support year, the earlier of the oldest supporting study in Open Targets and the GWAS Catalog year recorded by Minikel et al., fell in or before that year. We dated studies without a publication date, such as FinnGen release 12, by the public release of their summary statistics. For the 16,808 pairs, we restricted the analysis to support and phase entry documented by 2015 and counted support discovered later as absent. For the external data set, described next, support had to be documented by 2024, the start of its outcome window. Per phase transition, we fitted the GWAS-only model under each of the three gene assignments. As the external data set, we used the out-of-time set of PRIORITI^23^ (765 pairs), comparing pairs launched from 2025 with pairs discontinued or inactive after October 2024, adjusted for therapy area and pleiotropy, with Firth-penalised and leave-one-gene-out estimates as sensitivity analyses. We treated the label-free assignments as primary, since the L2G training set may include targets of drugs launched in 2025, and excluded pairs also present in our primary data set in a sensitivity analysis.

For trial-level analyses, we sourced trials from VirtualBiotech 2026^48^ and linked them to target– indication pairs through the drug, its recorded targets and the trial’s disease in ChEMBL. Each row combined a trial with one drug, one of its recorded targets and one disease term, so a trial could contribute several rows to a pair when it tested several drugs on the target or recorded several disease terms that mapped to the same indication. We clustered standard errors by gene and indication, and by gene only when the two-way variance of a reported estimate was not positive, which did not occur for any estimate reported here. Outcomes were a positive primary endpoint (40,100 rows with a known result) and stopping for lack of efficacy or for safety (80,490 rows). We assigned stopped trials to the first matching reason in a fixed order, first efficacy or safety, then commercial or operational reasons unrelated to the therapeutic hypothesis, then other reasons, and finally none recorded.

For the comparison with Minikel et al.^5^, we computed relative success as the crude ratio of launch rates with and without support, with Katz log confidence intervals. Their definition of GWAS support required an L2G share of at least 0.5 in Open Targets Genetics at the same similarity threshold. We grouped target-indication pairs into four sets, supported under both definitions, only under ours, only under theirs or under neither, and compared their launch rates. For pairs supported only under their definition, we recorded why ours did not support them, namely a study absent from Open Targets 26.06, curated top hits without a credible set, a credible set that no longer contained the gene, a measurement trait that our mapping did not link to a disease, or an L2G score below threshold. To test whether drug targets in the earlier L2G training set explained the difference, we removed all pairs whose gene was a drug-target example in that set (Open Targets Genetics v8 gold standard^5^) and recomputed relative success (**Supplementary Tab. 1**).

We did not adjust for multiple testing, because our analyses of modifiers form a descriptive set of estimates rather than a family of hypothesis tests^49^. Instead, we judged effect modification by its stability across analytical choices, following the idea of specification-curve analysis^50^, using the robustness criteria described below, and report P values only for joint Wald tests of non-linear or multi-term modifiers.

### Modifiers of the association

For each modifier separately, we fitted the GWAS-only model with the modifier as a main effect and an interaction with GWAS support. Continuous modifiers entered as restricted cubic splines to allow for non-linear associations, and we tested modification with a one-degree-of-freedom linear interaction per standard deviation. We report tertile and three-knot spline interactions as supplementary results. We chose the linear test as the primary test after inspecting the tertile results, without a formal pre-test for non-linearity. Approved mechanisms entered as a step (any versus none) and a slope per doubling, tested jointly.

We judged each modification against robustness criteria set before we fitted the first model of modifiers, guided by prior work that showed that findings depended on the cross-walk used to link genetic support to indications^4^. We refitted every estimate across 15 combinations of support thresholds, based on a grid of trait similarity (0.6, 0.7, 0.8, 0.9, 1.0) and gene-assignment confidence (L2G 0.3, 0.5, 0.7), and report the number of combinations in which the confidence interval excluded 1 in the primary direction, without a cut-off. A finding counted as robust if its direction held in all 15 combinations and its confidence interval excluded 1 across trait similarities 0.7 to 0.9 at L2G 0.5, and if its estimate under ChEMBL approval lay on the same side of 1.

We assigned indications to one of three settings by term rules applied in a fixed order. Risk factors included upstream usually quantitative states that predispose to disease rather than diagnosed illnesses (205 pairs, for example hyperlipidaemias, hypertension, tobacco use disorder). Established diseases included diagnosed, ongoing diseases, including all cancers (15,024 pairs). Acute conditions included time-limited events treated when they occur (1,204 pairs, for example stroke, sepsis, acute respiratory distress syndrome, COVID-19). The remaining 375 pairs, mostly signs and symptoms or indications without a clear setting, remained unassigned.

We further grouped indications by whether regulators accept a quantitative surrogate endpoint for approval, from the table of surrogate endpoints of the US Food and Drug Administration^51^. Our primary test compared indications with an accepted surrogate measured in the patient, such as HbA1c in diabetes, with indications without one, within established non-oncology disease. We did not exclude surrogate endpoints that had been validated by drugs acting on the target itself, because such a rule would select pairs by their outcome. Instead, we report a sensitivity analysis restricted to surrogate endpoints in use before 2017.

We obtained differential gene expression from disease contrasts in Expression Atlas^52^, and plasma protein associations from our prior work in UK Biobank^33^, which fitted Cox survival models of incident disease on Olink plasma proteins (43,240 participants), adjusted for age and sex, at a false discovery rate of 5%. Both entered a model that also adjusted for tractability, protein class, genetic constraint and tissue specificity. We summarised each target’s *cis*-acting genetic architecture by five quantitative trait locus (QTL) markers. From the GTEx v8^53^ eQTL credible sets in Open Targets 26.06, we took the largest variance in expression explained by a lead variant across tissues, the share of the 32 tissues with at least 200 samples that carried a *cis*-eQTL, and the spread of the explained variance across those tissues. From the GTEx v8 mashr results^54^, we took the number of the 49 tissues in which the lead independent eQTL was shared (local false sign rate below 0.05, same sign and at least half its maximal effect). From the pQTL credible sets in Open Targets 26.06, derived from the UK Biobank Pharma Proteomics Project^55^ and the INTERVAL study^56^, we took the share of plasma protein variance explained in *cis*. For targets measured only in INTERVAL, which contributed no *trans* credible sets, this share was 1 by construction. Each marker entered as a restricted cubic spline, and we report the odds ratio comparing the 75th with the 25th percentile.

### Drug x indication data set and drug mechanisms

We built drug x indication pairs from ChEMBL 36, with a maximum phase of at least 1 and a human target (32,101 pairs). A pair had GWAS support if any target of the drug was supported for the indication. For antibody-drug conjugates, the antigen defined the target. We adjusted for pleiotropy and therapy area and tested drug properties one at a time, with the same robustness criteria as in the analyses of modifiers. As sensitivity analyses, we weighted pairs for target annotation by phase and excluded oncology (**Supplementary Tab. 2**).

We classed antibody-drug conjugates, T-cell-redirecting therapies, checkpoint inhibitors and lineage-depleting antibodies by their mechanism of action. Because only one supported drug of these classes was approved, we report the 2 × 2 table, exact (Fisher) odds ratios within these drugs and within all other drugs, and, as the primary estimate of the interaction with inhibitory drugs, a Firth-penalised logistic regression, with two-way clustered standard errors as a sensitivity analysis.

### Comparison with Gentropy

We reproduced the estimand of the Gentropy analysis^29^, an unadjusted 2 x 2 odds ratio for ChEMBL approval under L2G assignment, and added in turn the GWAS-only covariates, two-way clustering by gene and indication, label-free gene assignment and leave-one-gene-out refits. Pleiotropy entered as log(count + 1) and its square, for the number of associated diseases (EFO terms) and of therapeutic areas, and we tested its modification of GWAS support with a two-degree-of-freedom Wald test.

### Discrimination

We fitted ridge logistic regression models and kept out-of-fold predictions from five folds grouped by gene, so that no drug target appeared in both training and test data. Features entered the model in blocks in a fixed order, starting with therapy area, then target biology, then tractability, location, redundancy, essentiality, annotation completeness and *cis*-QTL markers, and finally genetic support. The genetic block comprised GWAS, Mendelian, rare-variant, somatic and risk-factor-trait support in its four classes (for, against, untested, not testable), and pleiotropy. We quantified the unique contribution of a block as the loss in AUC when it was left out of the full model. We also assessed unpenalised logistic regression, least absolute shrinkage and selection operator (LASSO) regression and random forests (500 trees) using the same folds, with binary risk-factor-trait support and raw pleiotropy. The within-indication AUC compared launched with non-launched pairs of the same indication and fold. We estimated differences in AUC by a paired bootstrap over indications (2,000 resamples). To benchmark the genetic association score of Open Targets, which takes the maximum over diseases that reach the indication, we compared it with GWAS, Mendelian, rare-variant and somatic support. We obtained absolute launch probabilities and launches per 100 pairs by marginal standardisation from the reported models, with cluster bootstrap confidence intervals.

### Reproducibility

We generated all reported numbers in a single tagged run of the analysis code and checked them against a frozen register of values before submission. An independent re-implementation, written from the Methods text alone without access to our code, reproduced the analysis data set exactly and the main estimates closely. The code repository lists each analysis with its data set, outcome and model, together with the software versions and data releases. We refined model specifications during the analysis where initial specifications proved inadequate, for example in covariate coding and sample-size floors, and recorded each decision with its date in a log that is part of the code repository. We therefore present the analyses as descriptive rather than confirmatory.

### Use of AI tools

The authors designed the study, specified every analysis, made all analytical decisions and directed the implementation throughout. We used Claude (Anthropic), a large language model, through the Claude Code agent as an assistant to write, test and document the analysis code and the public code repository, and to help draft and edit the manuscript, figures and supplementary documents. A separate session of the same tool, without access to our code, performed the independent re-implementation described above. We checked every reported number against a frozen register and reproduced all results from a fresh copy of the repository. The authors take full responsibility for the content of this article.

## Data availability

All data analysed in this study are publicly available. Target-indication pairs and their development outcomes were taken from the data release of Minikel et al.^5^ (https://github.com/ericminikel/genetic_support), which derives from Citeline Pharmaprojects. We used only this public release. Human genetic evidence, gene assignments and clinical stages were taken from the Open Targets Platform release 26.06 (https://ftp.ebi.ac.uk/pub/databases/opentargets/platform/26.06/), and drug approvals and mechanisms from ChEMBL release 36 (https://ftp.ebi.ac.uk/pub/databases/chembl/ChEMBLdb/releases/chembl_36/). The independent outcome data set is the labelled PRIORITI data set from the supplementary material of Zhang et al.^23^, and trial-level outcomes are from the Virtual Biotech data release^48^. DEFA gene assignments are from Kohleick et al.^31^. Target annotations were taken from HGNC (https://www.genenames.org), the Human Protein Atlas v25 (https://www.proteinatlas.org), GTEx v8 (https://gtexportal.org) and OmniPath (https://omnipathdb.org). Summary results of associations between plasma proteins and disease onset in UK Biobank are deposited with the code repository. Exact versions, download dates and checksums of all inputs are listed in the code repository. All estimates reported are provided in Supplementary Tables 1-6 and the Source Data files. There are no restrictions on data availability.

## Code availability

The analysis code, together with the data needed to reproduce every result, is available to the editors and referees through a private Zenodo record. On publication, it will be released on GitHub under the MIT licence and archived on Zenodo with a DOI.

## Acknowledgements

The work was funded by the European Union (ERC, GenDrug, 101116072). Views and opinions expressed are however those of the author(s) only and do not necessarily reflect those of the European Union or the European Research Council. Neither the European Union nor the granting authority can be held responsible for them. The funders had no role in study design, data collection and analysis, decision to publish or preparation of the manuscript.

## Author contributions

M.P. conceived and designed the study, performed the analyses and wrote the manuscript. L.K. developed DEFA and the label-free gene assignment and contributed to the analyses. Both authors interpreted the results and revised the manuscript.

## Additional information

Supplementary Information is available for this paper. Correspondence and requests for materials should be addressed to Maik Pietzner.

## Extended Data figure legends

**Extended Data Fig. 1 Launch rate by confidence of the gene assignment.** Launch rate of GWAS-supported pairs by quartile of assignment probability for DEFA (897 pairs) and L2G (773 pairs), with quartiles defined within each assignment. Points show launch rates, lines Wilson 95% confidence intervals, and numbers the pairs per quartile.

**Extended Data Fig. 2 *Cis*-QTL markers of the target and launch.** ORs for launch comparing the 75th with the 25th percentile of five markers of the target’s *cis*-acting genetic architecture (Methods), from inverse-probability-weighted models. Lines show 95% confidence intervals; none excluded 1.

**Extended Data Fig. 3 Predictors selected by LASSO.** All non-zero coefficients of the LASSO model (out-of-fold AUC 0.661), grouped by block (genetic support, target biology, tractability, *cis*-QTL markers and indication), with genetic support terms in the colour of their evidence type (cyan, Mendelian; khaki, burden test; brown, somatic; coral, GWAS) and all other terms in grey, negative coefficients in a lighter shade. Continuous predictors entered as spline terms.

