## Supplementary Note and Figures for "Human genetic support improves the odds of drug success but poorly predicts it"

#### Supplementary Note 1: Why most launched drugs lacked GWAS support

Most target-indication pairs that yielded a launched drug had no GWAS support. To understand why, we assigned each unsupported pair to the first matching reason in a fixed order and compared how often each reason occurred among launched and among not-launched pairs (ratio of shares, where values above 1 mark reasons that were more common among launches) (**Supplementary Tab. 6**). Two thirds of unsupported launches (66%) had an identifiable reason. The remaining third was unexplained, a smaller share than among pairs that did not launch (34% compared with 45%).

| **Reason (first match)** | **Share of unsupported launches** | **Ratio of shares, launched vs not launched** |
|---|---|---|
| Other genetic evidence (Mendelian, rare-variant or somatic support, or concordant risk-factor-trait support) | 17% | 1.73 |
| No usable GWAS for the indication | 18% | 1.77 |
| Target invisible to GWAS (major histocompatibility complex or X chromosome) | 3% | 0.68 |
| GWAS signal below the support thresholds | 8% | 1.09 |
| Symptomatic or replacement indication | 18% | 1.11 |
| Housekeeping target | 3% | 0.45 |
| Unexplained | 34% | 0.74 |

Two reasons were more common among launches. Launched drugs without GWAS support more often had other genetic evidence for the same indication (category 1). They also more often treated indications without a usable GWAS (category 2), although this did not hold under ChEMBL approval (ratio of shares: 0.97). We note that reasons were not mutually exclusive. When we counted every reason that applied to a pair, rather than only the first, a third of unsupported launches were for indications usually treated symptomatically, such as pain or depression, or by replacing a missing function, such as hormone or enzyme deficiencies, compared with a quarter of pairs that did not launch (33% compared with 25%). We assigned this category from the therapy area and the indication term rather than from the drug. For symptomatic treatments, the target need not be causal for the disease, so genetic support is not necessarily expected, whereas for replacement therapies the target is often the causal gene. This category coincided with other genetic evidence for 133 pairs and with indications lacking a usable GWAS for 874. Among unexplained pairs, 31% of launched and 27% of not-launched pairs involved targets of drug classes that use their target only to reach particular cells, such as antibody-drug conjugates, rising to 46% among launched oncology pairs.

An example to better understand the categories were anti-TNF antibodies. Of 15 launched anti-TNF pairs without GWAS support, 4 had other genetic evidence and 9 were hard to infer from GWAS, because *TNF* lies in the major histocompatibility complex, a region where variant-to-gene assignment is unreliable. Five of the 15, among them rheumatoid arthritis and Crohn's disease, would have been supported only through the 2024 partner support (Results), which is not considered here. In both versions of partner support, the links referred to proteins of the TNF signalling complex, with *TNIP1* among them in every indication (**Supplementary Tab. 6**).

Most unsupported launches therefore reflect gaps in GWAS coverage, such as indications without a usable GWAS or targets in the major histocompatibility complex, or indications usually treated symptomatically, rather than a failure of genetic evidence where it exists.

### Extended Data Figures

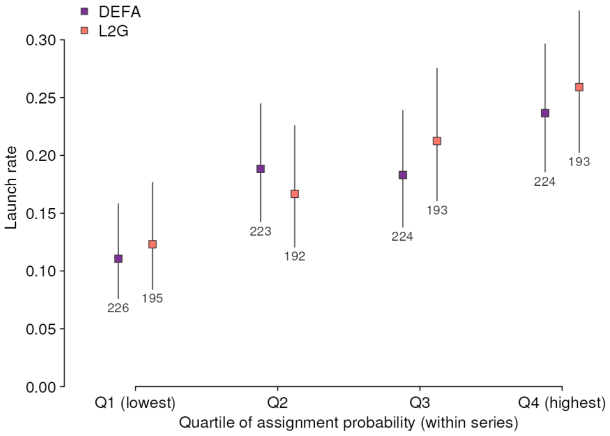


**Extended Data Fig. 1 | Launch rate by confidence of the gene assignment.** Launch rate of GWAS-supported pairs by quartile of assignment probability for DEFA (897 pairs) and L2G (773 pairs), with quartiles defined within each assignment. Points show launch rates, lines Wilson 95% confidence intervals, and numbers the pairs per quartile.

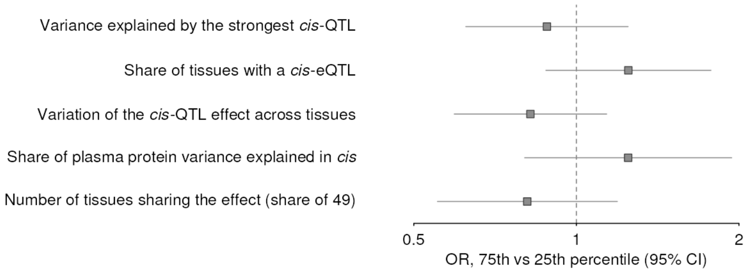


**Extended Data Fig. 2 | *Cis*-QTL markers of the target and launch.** ORs for launch comparing the 75th with the 25th percentile of five markers of the target's *cis*-acting genetic architecture (Methods), from inverse-probability-weighted models. Lines show 95% confidence intervals; none excluded 1.

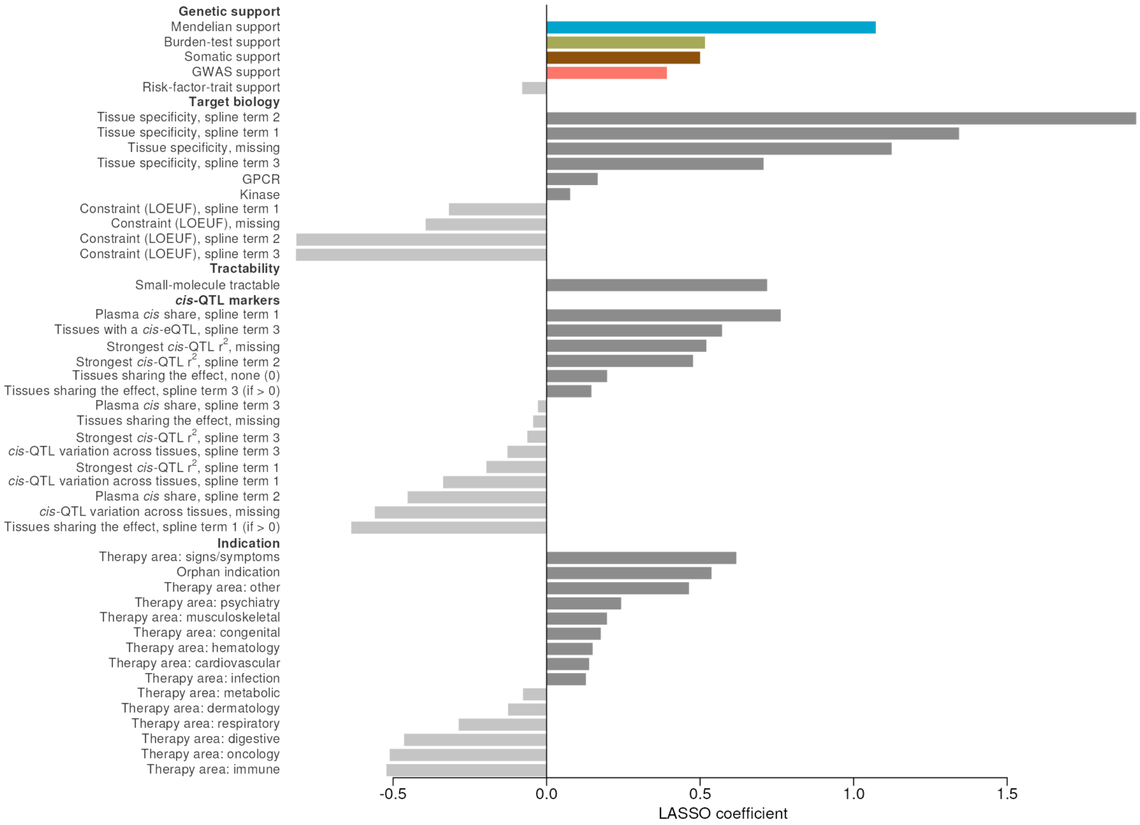


**Extended Data Fig. 3 | Predictors selected by LASSO.** All non-zero coefficients of the LASSO model (out-of-fold AUC 0.661), grouped by block (genetic support, target biology, tractability, *cis*-QTL markers and indication), with genetic support terms in the colour of their evidence type (cyan, Mendelian; khaki, burden test; brown, somatic; coral, GWAS) and all other terms in grey, negative coefficients in a lighter shade. Continuous predictors entered as spline terms.
